# Effective contact tracing can halve mass testing frequency needed to control respiratory virus pandemics

**DOI:** 10.1101/2023.10.12.23296969

**Authors:** Oraya Srimokla, James Petrie

## Abstract

Mass testing is a promising strategy to control future respiratory virus pandemics, however, it is limited by the availability of testing capacity. The aim of this work is to predict the effectiveness of contact tracing when used with mass testing, and to what extent contact tracing can reduce the required number of tests. We estimate the effect of contact tracing and mass testing for controlling a variety of respiratory virus pandemics. To do this, we use a branching model with individuals whose infectiousness and probability of testing positive depend on simulated viral load trajectories. We find that the addition of contact tracing is most useful when mass testing is done frequently enough to detect most infections, but cannot be done frequently enough to reliably isolate cases before they infect others. Our results show that the addition of effective contact tracing can prevent the same number of transmissions as doubling the mass testing frequency.

## 1 Background

Respiratory virus pandemics like SARS-CoV-2 and the 1918 Influenza pandemic have caused massive harm, and more pandemics are likely in the future [21]. Despite recent efforts to control disease threats, significant challenges still exist in improving global pandemic preparedness [38, 9, 8]. Several researchers proposed mass testing (frequent testing of most individuals) as a method to control Covid-19 [36, 20, 17, 26, 33, 18, 6]. Relatively small-scale or short-term use of mass testing was implemented in several locations [35, 28, 34, 16, 13], along with extensive use in China [41]. Compared to other non-pharmaceutical interventions like social distancing, mass testing bears less of a burden than imposing widespread social distancing [32].

In Petrie et al. [30], we simulated viral load trajectories of potential pathogens to investigate how effective mass testing could be against future respiratory virus pandemics. Frequent testing of most of the population and isolation of positives was shown to be able to control respiratory virus pandemics if there is sufficient adherence to testing and isolation. The model used in Petrie et al. is based on how frequent testing reduced transmissions from a single infected individual. While the single-individual model is able to predict population-level effects that are independent of other individuals like scheduled testing or masking, it is unable to incorporate more complex non-pharmaceutical interventions (NPIs) like contact tracing.

Contact tracing is a tool used to mitigate disease spread that involves identifying the contacts of the infected people and quarantining them before they can infect others [37]. Modelling by Fraser et al. demonstrates that diseases with low presymptomatic or asymptomatic transmission can be controlled by contact tracing if there is high adherence to tracing and isolation [11]. However, diseases like SARS-CoV-2 and HIV with significant presymptomatic or asymptomatic transmission are more difficult to control and require additional interventions to contain [11]. Digital contact tracing was proposed as a faster alternative to manual contact tracing [9], however, the effectiveness of digital contact tracing is more sensitive to public adherence at multiple steps in the notification process [22].

Combining contact tracing with mass testing is a promising approach to reduce transmissions without using significantly more tests. This is important because testing capacity is both expensive and limited, and fast-spreading pathogens might not be sufficiently controlled by mass testing alone. Modelling studies showed that a combination of contact tracing and mass testing could potentially control COVID-19 transmission in the United Kingdom [23]. Similar findings were also observed by Zhou et al. in China [41] and Cheng et al. in Hong Kong [5]. Armbruster and Brandeau analysed the cost-effectiveness of contact tracing vs. screening for reducing the prevalence of endemic disease [2]. However, prior to this work, there was limited research on the combined use of mass testing and contact tracing for controlling future respiratory virus pandemics.

### 1.1 Aims & Objectives

Our aim is to evaluate how much the addition of contact tracing can improve a mass testing strategy. We investigate how this impact depends on the pathogen (time to peak viral load, *R*_0_, and timing of detection of symptoms) and the mass testing strategy (testing frequency, test result delay time, and test sensitivity). To identify if a combination of mass testing and contact tracing is effective for controlling future outbreaks, we evaluate these methods against pathogens similar to previous epidemics (1918 Influenza, SARS-CoV-1, and SARS-CoV-2) as well as a range of other pathogens (characterised by *R*_0_ and time to peak viral load).

## 2 Methods

### 2.1 Model Development

We developed a stochastic branching model to simulate how outbreaks are affected by frequent testing and contact tracing. In our branching model, we initialise the number of infected cases to start an outbreak and update the infected sub-population every hour depending on testing, symptom detection, contact tracing, and transmissions. We simulate many rounds of outbreaks and calculate the effective reproduction number, *R*_*e*_ [27], by averaging the number of people infected for each infector. Similar to the model used in Petrie et al. [30], viral load trajectories are used to mechanistically model infectiousness and test sensitivity over time. This is further described below.

#### 2.1.1 Viral Load, Infectiousness, and Test Sensitivity

Trajectories of log viral loads for acute respiratory infections are modelled using piecewise linear curves [16, 4, 7]. Viral load trajectories of log viral load, log_10_(*V* (*t*)), are calculated using characteristics of pathogens including their peak viral load, *V*_*p*_, the time from infection to peak viral load, *τ*_*p*_, and the time from peak viral load to recovery, *τ*_*r*_. The initial viral load when infected, *V*_*o*_, is set to 3*\**10^*−*3^ from the initial infection of one viral particle and about 300ml of respiratory fluid [19]. This is shown in Equation 1.

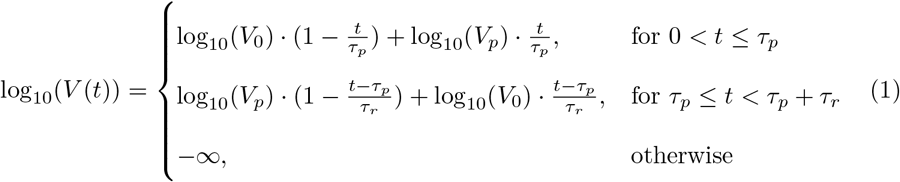

The test sensitivity (probability that a sample from an infected person is detected as positive), *S*_*V*_ (*V*), is calculated using the viral load and is shown in Equation 2. The equation uses the viral load at which 50% of samples from infected people are positive, LOD_50_, the width of the intermediate regions, *k*, and the peak test sensitivity, *S*_*max*_. For PCR tests, we set *k* = 6 so that the distance between 5% and 95% sensitivity is about a multiple of 10 as in [40], *S*_*max*_ is set to 99.5% to account for sample mishandling, and LOD_50_ = 10^2^ copies/ml [40].

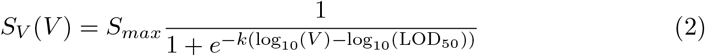

To estimate the relationship between infectiousness and viral load, the saturation model from Ke et al. is used [15]. The expected transmission rate, depending on viral load, *T*_*V*_ (*V*), is parameterised using the average number of contacts per hour *N*_*c*_ = 13*/*24 [25], shape parameter, *h*, parameter controlling maximum infectiousness per interaction, *θ*, and the viral load where half of peak infectiousness is reached, *K*_*m*_ = 8.9 *\** 10^6^*copies/ml*. This is shown in Equation 3.

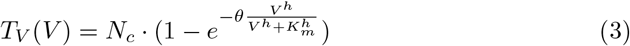

Using the infectiousness model (Equation 3) and the viral load over time (Equation 1), the expected number of transmissions from each infected person is calculated every hour. The branching model is discrete (an integer number of people are infected) and stochastic, so we make the simplifying assumption that transmission events are independent and sample the number of hourly transmissions from each person using a Poisson distribution. We set the Poisson rate parameter (and therefore the mean of the sampled distribution) to the expected number of transmissions, which is *λ*(*t*) = *T*_*V*_ (*V* (*t*)) if the infected person is not in quarantine or isolation (as discussed in Section 2.1.2).

#### 2.1.2 Contact Tracing and Frequent Testing

In our model, infected individuals can either be undetected, symptomatic, quarantined, or isolated, and they move between these states as shown in Figure 1. All infected individuals start as undetected and their state is updated every hour depending on test results, noticing symptoms, or if they are contact traced. A disease-specific fraction of infected individuals notice symptoms and request additional testing because of this, as shown in Table 1. For individuals that notice symptoms, the time that symptoms develop is set relative to the time of peak viral load by the disease-specific parameter, *timeFromPeakToSymptoms* shown in Table 1.

**Table 1:** Disease Specific Parameters Used in the Branching Model.

| Respiratory Pandemic | Parameter | Value | Reference |
| --- | --- | --- | --- |
| 1918 Influenza | Probability Develop Symptoms | 0.693 <sup>3</sup> | [4] |
| 1918 Influenza | Probability Testing Once Symptomatic | 0.200 | Estimated <sup>1</sup> |
| 1918 Influenza | Time Peak Viral Load to Symptoms | -2 | [29] |
| 1918 Influenza | Days from Infection to Peak Viral Load | 4 | [24] |
| 1918 Influenza | R0 | 2 | [29] |
| SARS-CoV-1 | Probability Develop Symptoms | 0.925 | [39] |
| SARS-CoV-1 | Probability Testing Once Symptomatic | 0.800 | Estimated <sup>2</sup> |
| SARS-CoV-1 | Time Peak Viral Load to Symptoms | -6 | [29] |
| SARS-CoV-1 | Days from Infection to Peak Viral Load | 10.5 | [29] |
| SARS-CoV-1 | R0 | 2.4 | [29] |
| SARS-CoV-2 | Probability Develop Symptoms | 0.800 | [14] |
| SARS-CoV-2 | Probability Testing Once Symptomatic | 0.200 | Estimated <sup>1</sup> |
| SARS-CoV-2 | Time Peak Viral Load to Symptoms | 0 | [29] |
| SARS-CoV-2 | Days from Infection to Peak Viral Load | 5 | [29] |
| SARS-CoV-2 | R0 | 2.5 | [29] |

<sup>1</sup>0.20 was selected as an initial value. Alternative values are tested and can be found in the Appendix A
<sup>2</sup>0.80 was selected as an initial value since the probability of developing symptoms after testing positive should be greater than hospitalisation values for SARS-CoV-1 [29]. Alternative values are tested and can be found in the Appendix A
<sup>3</sup>Value used is estimated based on seasonal Influenza

**Fig. 1:**
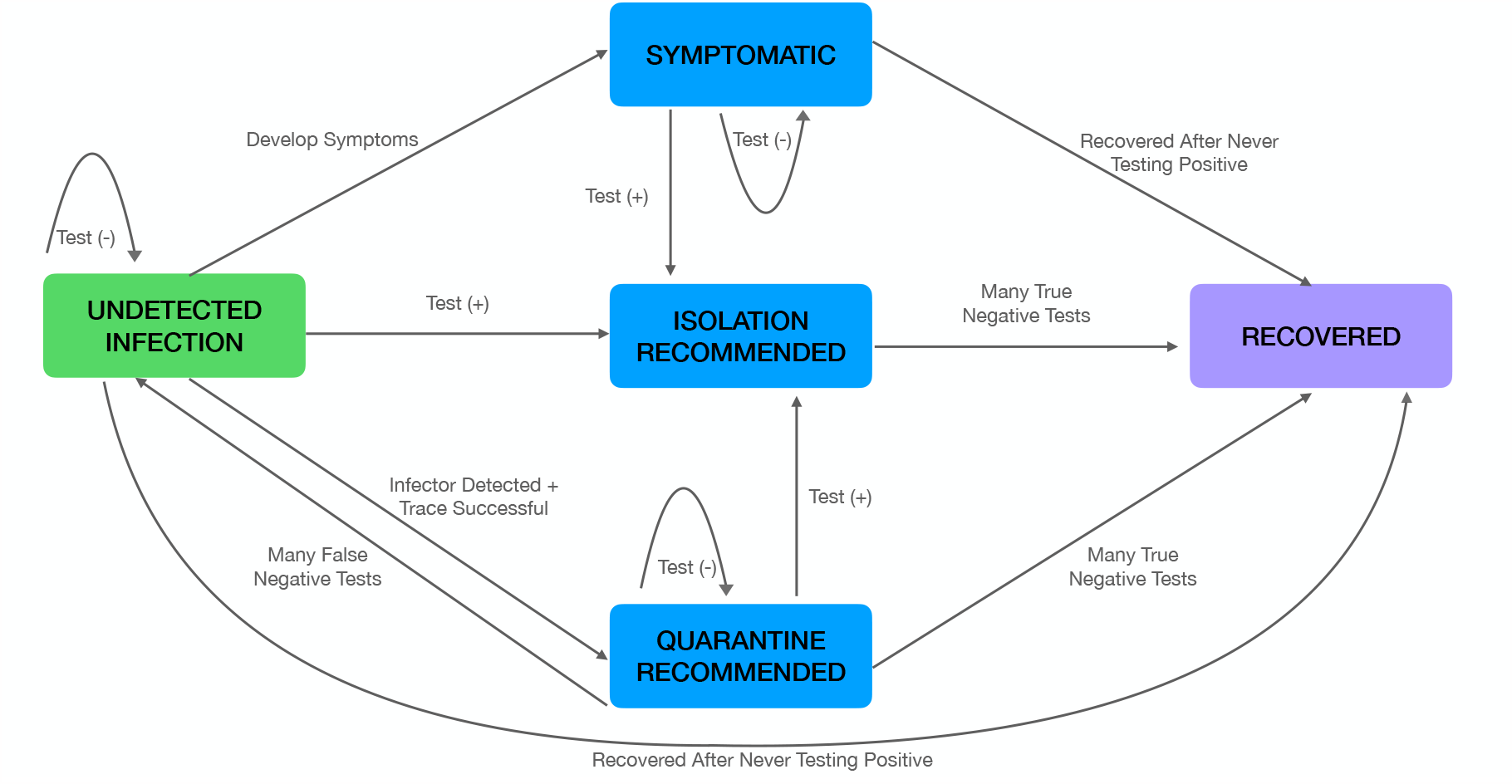
Contact Tracing Model Transmission States. The state model shows how an infected individual moves between undetected, quarantined, isolated, symptomatic, and recovered states depending on testing, symptoms, and contact tracing. The transmission rate from individuals is modified depending on which state they are in. We start with an undetected infection state and either remain as an undetected infection if the individual continues to test negative or transition to the symptomatic, isolated, quarantined, or recovered state. The infected individual is moved to quarantine state when their infector has been detected and contact tracing has been successful. At this state, the transmission is reduced by the effectiveness of quarantine value. The same transmission reduction is also shown when an infected individual tests positive and moves to isolation. Once isolated and having tested negative multiple times, the individual is then moved to the recovered state. If an infected individual becomes symptomatic and continues to test negative, they are transitioned to the recovered state after never testing positive. From the quarantined state, they are moved back to an undetected infection state if many false negative tests are produced or the recovered state if many true negative tests are produced.

Individuals that have received a positive test are recommended to isolate, and their transmission rate is reduced by the isolation effectiveness (80% for most of this manuscript and other values are shown in Figure B4). Individuals who are contact traced but have not yet tested positive are recommended to quarantine and their transmission rate is similarly reduced. A portion of the contacts of people who have tested positive are successfully contact traced and recommended to quarantine, as set by the *FracTraced* parameter. Contacts that are traced are notified 24 hours after the person that infected them receives a positive test result, similar to either fast manual contact tracing or a typical notification delay for digital contact tracing [12].

Individuals are recommended to test every *TestPeriod* days as part of the mass testing strategy, and they are also recommended to test daily if they are contact traced or symptomatic. The fraction of individuals that adhere to any testing recommendations is set to 90% for the remainder of this manuscript. When an individual is tested, the probability that the test is positive is given by Equation 2 for the viral load at the time of the test. The individual learns the result of the test *TestDelay* hours later, which is set to 10 hours for the remainder of this manuscript.

### 2.2 Control of Example Pathogens

To investigate the usefulness of mass testing and contact tracing against possible pandemic pathogens, we start by evaluating these strategies with pathogen-specific characteristics chosen to match 1918 Influenza, SARS-CoV-1, and SARS-CoV-2. Using the disease characteristics shown in Table 1, the branching model is run for each scenario. The effectiveness of these interventions is measured in terms of the reduction in the effective reproduction number, *R*_*e*_. We estimate *R*_*e*_ based on the average number of transmissions from each infected person who has recovered at the time the simulation ends. We also compute bootstrapped 95% confidence intervals for these values using the Basic method from the R library ‘boot’ [1].

## 3 Model Results

We analyse the effectiveness of the combination of mass testing and contact tracing interventions in terms of *R*_*e*_. Variations of the testing period and the fraction of contacts traced are used to examine the effect of these interventions for different pathogens. In Figure 2, *R*_*e*_ is shown as a result of different testing periods and fractions of contacts traced when applied to the 1918 Influenza, SARS-CoV-1, and SARS-CoV-2 pandemics.

**Fig. 2:**
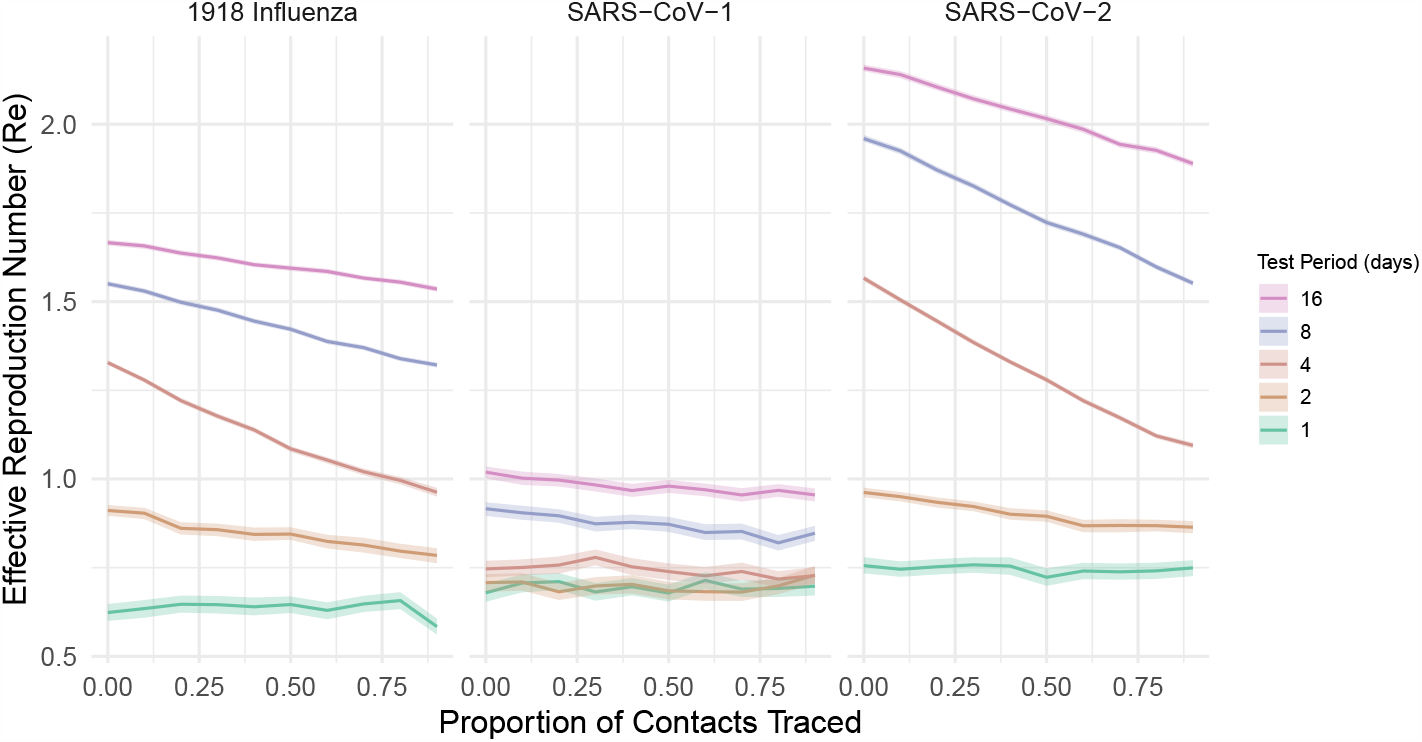
*R*_*e*_ Depending on Testing Period and Proportion of Contacts Traced. *R*_*e*_ for the 1918 Influenza pandemic (left) is computed using pathogen-specific parameters shown in Table 1, a contact tracing delay of 24 hours, and an adherence to isolation of 0.90. Similarly, *R*_*e*_ for SARS-CoV-1 is shown (middle) and SARS-CoV-2 (right). All plots show *R*_*e*_ as a result of variations in contact tracing and testing period. With frequent testing, the increase in contact tracing does not significantly reduce transmission (shown by having a flatter trend line) since mass testing is able to control transmission on its own. With less frequent testing, contact tracing has more influence in reducing transmission (shown by a steeper slope). For SARS-CoV-1, *R*_*e*_ is close to 1 even with infrequent testing because a significant fraction of people get tested when they are symptomatic and as a result, isolate before infecting others. Appendix A analyses the same scenarios when a smaller fraction of people seek testing after developing symptoms.

For the 1918 Influenza pandemic (left side of Figure 2), there is a decline in *R*_*e*_ with an increasing proportion of contacts traced for test periods of 2 to 16 days. A steeper slope means there is a greater marginal benefit of additional contact tracing. The slope is steepest with a test period of 4 days. With a larger test period fewer index cases cases are found through mass testing and symptomatic reporting is not consistent enough to counteract this. With fewer index cases detected, fewer contacts are able to be traced, which leads to a relatively flatter slope for test periods of 8 and 16 days. Conversely, when testing is very frequent relative to the duration of infection, most cases are already detected before they infect others. This is why the slope is flat when using daily testing for 1918 Influenza. For the scenario with mass testing every 4 days, the addition of very effective contact tracing reduces *R*_*e*_ from 1.3 to 0.95, which would be sufficient to bring the outbreak under control without social distancing. This reduction in *R*_*e*_ is roughly equivalent to the benefit from testing every 2 days instead of every 4 days.

For SARS-CoV-1 (middle of Figure 2), we see an *R*_*e*_ value close to 1 even with infrequent testing since a higher proportion of people develop symptoms, get tested, and isolate before they become significantly infectious. Because the disease is already well controlled, the benefit of mass testing and contact tracing is more limited. In Appendix A we evaluate a version of SARS-CoV-1 with a lower fraction of people testing because of symptoms, and find higher initial values of *R*_*e*_ and more benefit from mass testing and contact tracing.

For SARS-CoV-2 (right side of Figure 2), we observe patterns similar to the 1918 Influenza example: the addition of contact tracing has the largest benefit when testing every 4 days, and there is no noticeable benefit to adding contact tracing when testing daily. Differently from 1918 Influenza, *R*_*e*_ is higher with infrequent testing and no contact tracing (2.2 instead of 1.7), and the slope is more steep when testing every 16 days. One reason for the steeper slope is that the duration of infection for SARS-CoV-2 is longer than Influenza (Table 1), so more cases are detected and can have their contacts traced.

Further exploration of Figure 2 with variations in the probability a person tests once symptomatic and adherence to isolation is shown in Appendix A and Appendix B.

To evaluate a combination of mass testing and contact tracing for other pathogens, Figure 3 shows how *R*_*e*_ changes with 6 different values for the time from infection to peak viral load and a range of test periods between daily and monthly testing. In each plot, the improvement gained by adding very effective contact tracing is quantified by the change in *R*_*e*_ between the upper (0% traced) and lower (90% traced) lines. More modest improvements in contact tracing are roughly a proportional distance between these lines, as demonstrated by the 50% line in the middle.

**Fig. 3:**
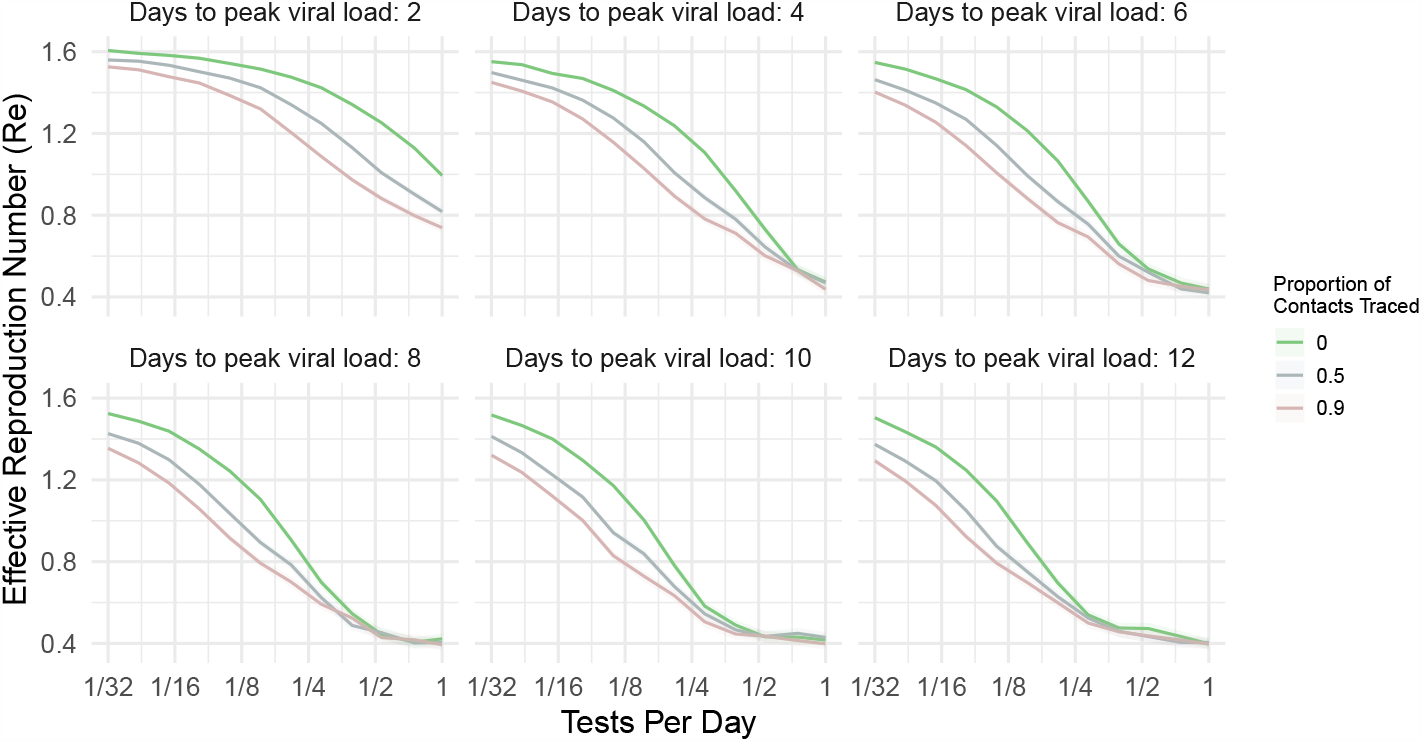
*R*_*e*_ Depending on Testing Frequency With and Without Contact Tracing. Subplots are generated for potential pathogens with viral load trajectories that reach peak viral load 2 to 12 days after infection. More frequent testing reduces *R*_*e*_ until the benefit saturates because most infections have already been found. The addition of effective contact tracing significantly reduces *R*_*e*_ if mass testing is frequent enough to find most cases, but not frequent enough to reliably isolate cases before they infect others. Parameters used for the analysis are: probability of noticing symptoms = 0.50, contact tracing delay = 24 (hours), probability of testing because of noticing symptoms = 0.5, time from peak viral load to symptoms = -1 days, and *R*_0_ = 2.

In all plots, more frequent testing reduces *R*_*e*_ until the marginal benefit of increased testing is saturated – roughly when the testing period is 4 times shorter than the time to reach peak viral load. After this point, the benefit of additional testing or contact tracing is limited because almost all infected people who adhere to testing are already found before they are significantly infectious. Similarly, when testing is very infrequent relative to the time to peak viral load, the change in *R*_*e*_ from further reducing the testing frequency also saturates. Without mass testing in this scenario, the fraction of infected individuals found through symptomatic testing is small, and these detected individuals are responsible for a much smaller fraction of transmissions (because they are recommended to isolate after testing positive). The small fraction of index cases detected is why the benefit of contact tracing is much smaller when mass testing is infrequent for the pathogen with 2 days to peak viral load in Figure 3.

When mass testing is frequent enough to detect most infections, but not frequent enough to prevent most transmissions, effective contact tracing can substantially reduce the number of expected transmissions. For moderately frequent testing of the pathogens examined in Figure 3, the reduction in *R*_*e*_ from effective contact tracing roughly matches the reduction caused by doubling the testing frequency. For example, for the pathogen with 6 days to peak viral load in Figure 3, adding highly effective contact tracing when mass testing every 8 days reduces *R*_*e*_ from 1.3 to 0.95. This is roughly the same reduction achieved by testing every 4 days without effective contact tracing. Similarly, the addition of moderately effective contact tracing (50% of contacts) with moderately frequent mass testing roughly reduces *R*_*e*_ by the same amount as a 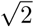 increase in testing frequency (half the distance between labelled ticks on the log scale).

## 4 Discussion

Results from our viral-load-based branching model show combining contact tracing with mass testing can significantly reduce *R*_*e*_ for a variety of respiratory viruses. Unlike when using contact tracing by itself, as modelled by Fraser et al. [11], contact tracing combined with mass testing is much less dependent on symptom-based detection of cases.

We find that the addition of contact tracing is most useful when mass testing is done frequently enough to detect most infections, but not frequently enough to consistently isolate cases before they infect others (roughly when *TestPeriod* = *TimeToPeak*). In this case, the addition of moderately effective contact tracing (reaching 50% of contacts) causes the same reduction in *R*_*e*_ as a 40% increase in testing frequency. Moderately effective contact tracing could potentially be achieved by focusing tracing on family members or contacts with very long exposure [10]. With highly effective contact tracing (90% of contacts found), the reduction in *R*_*e*_ is the same as doubling the frequency of mass testing. This means that the addition of effective contact tracing could be used to halve the number of tests – and therefore the cost – of mass testing while maintaining the same control of a pandemic.

If the test period is significantly longer than the duration of infection, the reduction in *R*_*e*_ caused by adding contact tracing is more modest and depends more strongly on symptomatic detection. Similarly, if the test period is significantly shorter than the duration of infection, most cases are already found early in infection without requiring contact tracing. The change in *R*_*e*_ could also be smaller if adherence to quarantine (variations of this value are shown in Appendix B4) or testing is low, or if contact tracing is slower than modelled here.

Even in scenarios where the expected reduction in *R*_*e*_ from contact tracing is relatively small, it could still be highly cost effective to implement if the overall disease prevalence is low. This is because the cost of contact tracing is proportional to the number of people that are infected, which could vary over many orders of magnitude, while the cost of social distancing or mass testing is roughly constant with prevalence [31]. For SARS-CoV-2, Brauner et al. estimated that school and university closures caused a 38% reduction in *R*_*e*_ [3]. Even partially reducing the need for these closures, e.g. by reducing *R*_*e*_ by 5% with contact tracing, could have a large positive impact.

Overall, our model shows that a combination of contact tracing and mass testing can effectively control a variety of respiratory virus pandemics. With a mass testing period roughly the same as the time to reach peak viral load, the addition of effective contact tracing results in the same reduction in transmissions as doubling the number of tests used.

## Data Availability

All code and data for the present study are available upon reasonable request to the authors

### Appendix A Influence of Probability of Testing After Noticing Symptoms

In this section, the probability that a person gets tested after noticing symptoms is varied. The dependence of *R*_*e*_ on the proportion of contacts traced and the testing period is visualised.

From Figure A1, we reduce the probability that a person gets tested after noticing symptoms to 0.10. When compared to Figure A2 with a probability of 0.50 for Influenza, SARS-CoV-1, and SARS-CoV-2, we observe a higher transmission magnitude with a lower probability that a person tests once they are symptomatic. As we increase the probability a person tests once they are symptomatic from 0.10 to 0.90, we see slightly flatter slopes for the 1918 Influenza for test periods of 2, 4, 8, and 16 hours while flat slopes are observed for a testing period of 1 day. In this case, mass testing and contact tracing have more impact in reducing transmission when the probability a person gets tested once symptomatic is low. Flatter slopes are observed for SARS-CoV-1 as we increase this probability. This shows that with a lower probability a person tests once they are symptomatic for SARS-CoV-1, mass testing and contact tracing have more of an impact in reducing transmission (shown by steeper slopes when contact tracing is increased) for more infrequent testing periods of 8 and 16 days. In this scenario, we see that testing periods of 1, 2, and 4 days are frequent enough to bring transmission to a controllable level and the increase in contact tracing does not have a significant impact on the transmission. We observe that with a probability a person tests after noticing symptoms of 0.90, the transmission is already at a controllable level even with infrequent testing since a higher proportion of infectious people are found with increased testing once symptomatic. For SARS-CoV-2, we see that with an increase in the probability a person tests once they are symptomatic from 0.10 to 0.90, the magnitude of transmission decreases with a testing period of 4, 8, and 16 days while staying the same for testing periods of 1 and 2 days. Additionally, when increasing the probability of testing once symptomatic, the slopes for test periods of 4, 8, and 16 days become steeper. Under these conditions with infrequent testing periods, mass testing and contact tracing have more impact in reducing transmissions when more people are testing once symptomatic. Flatter slopes are observed for testing periods of 1 and 2 days for SARS-CoV-2 since frequent mass testing is able to reduce transmission to a controllable level on its own.

When comparing these three respiratory pandemics, we see how different strategies can be designed for different pathogens to bring their transmission to a controllable level. All three pathogens have different disease characteristics such as the probability of being symptomatic after infection, the time to peak viral load, and the time from symptoms to peak viral load. Interventions can be designed based on disease and behavioural characteristics to more effectively implement and reduce disease transmission.

### Appendix B Influence of Isolation Effectiveness

We simulate different scenarios with different fractions of proper isolation and estimate the *R*_*e*_ as a result. We use a combination of testing periods of 2 and 8, and the proportions of contacts traced of 0.10 and 0.50 for our example pathogens with variations in the effectiveness of isolation and quarantine.

From Figure B4, we see that with a more frequent testing period and a higher proportion of contacts traced, the greater the transmission is reduced with an increase in the fraction of contacts isolated. With a lower fraction of contacts isolated less than 0.10, we see that more frequent testing is able to slightly reduce transmission for 1918 Influenza and SARS-CoV-2 while the increase in contact tracing was not able to significantly decrease transmission. As the fraction of contacts isolated increases, we notice that there is a greater reduction in transmission with a higher proportion of contact tracing for a test period of 8 days for 1918 Influenza and SARS-CoV-2. For the 1918 Influenza, a fraction of contacts isolated of around 0.70, contacts traced of 0.50, and a testing period of 2 days is able to reduce transmission to a controllable level with an *R*_*e*_ less than 1. This is similarly observed for a fraction of isolation around 0.75 for a testing period of 2 days and traction of contacts traced of 0.10. For SARS-CoV-1, a controllable transmission level is achieved using a testing period of 2 days with around 0.62 fraction of contacts isolated, and 0.50 or 0.10 proportions of contacts traced. This is similarly observed when having around 0.75 fraction of contact isolated with a testing period of 8 days and 0.50 or 0.10 proportions of contacts traced. For SARS-CoV-1, we see that the increase in contacts traced does not make a difference in reducing transmission for the same testing period and the effectiveness of isolation and quarantine. This is also similarly observed in SARS-CoV-2 for a testing period of 2 days. For SARS-CoV-2, we see that with an increase in the effectiveness of isolation and quarantine, an increase in contact tracing is able to reduce transmission more if people are effectively isolating and quarantining. For pathogens like 1918 Influenza and SARS-CoV-2, we see that even with a 0.30 proportion of people isolating and quarantining, contact tracing can be used to reduce transmission with infrequent testing. When paired with mass testing, transmission can be further reduced even with a low proportion of people isolating and quarantining.

**Fig. A1:**
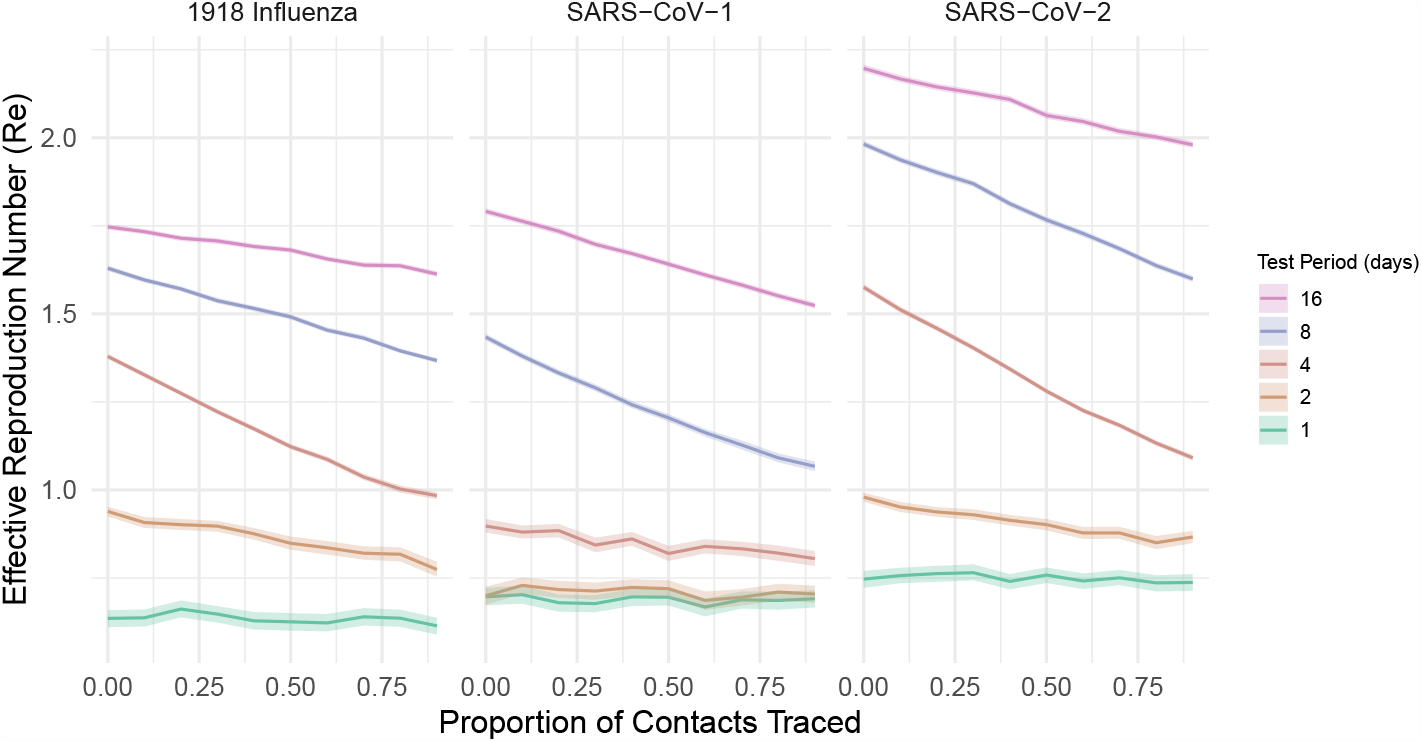
Re Depending on Testing Period and Proportion of Contacts Traced with Probability a Person Tests After Noticing Symptoms = 0.10. The *R*_*e*_ for the 1918 Influenza pandemic (left) is created using pathogen-specific characteristics shown in Table 1 (excluding the probability of testing once symptomatic), a contact tracing delay of 24 hours, adherence to isolation of 0.90, and the probability a person gets tested after being symptomatic is 0.10 (note that the probability of developing symptoms for Influenza is 0.693, SARS-CoV-1 is 0.925, and SARS-CoV-2 is 0.800). Similarly, the *R*_*e*_ for SARS-CoV-1 is shown (middle) and SARS-CoV-2 (right). Both plots show various trends as a result of variations in testing period.

**Fig. A2:**
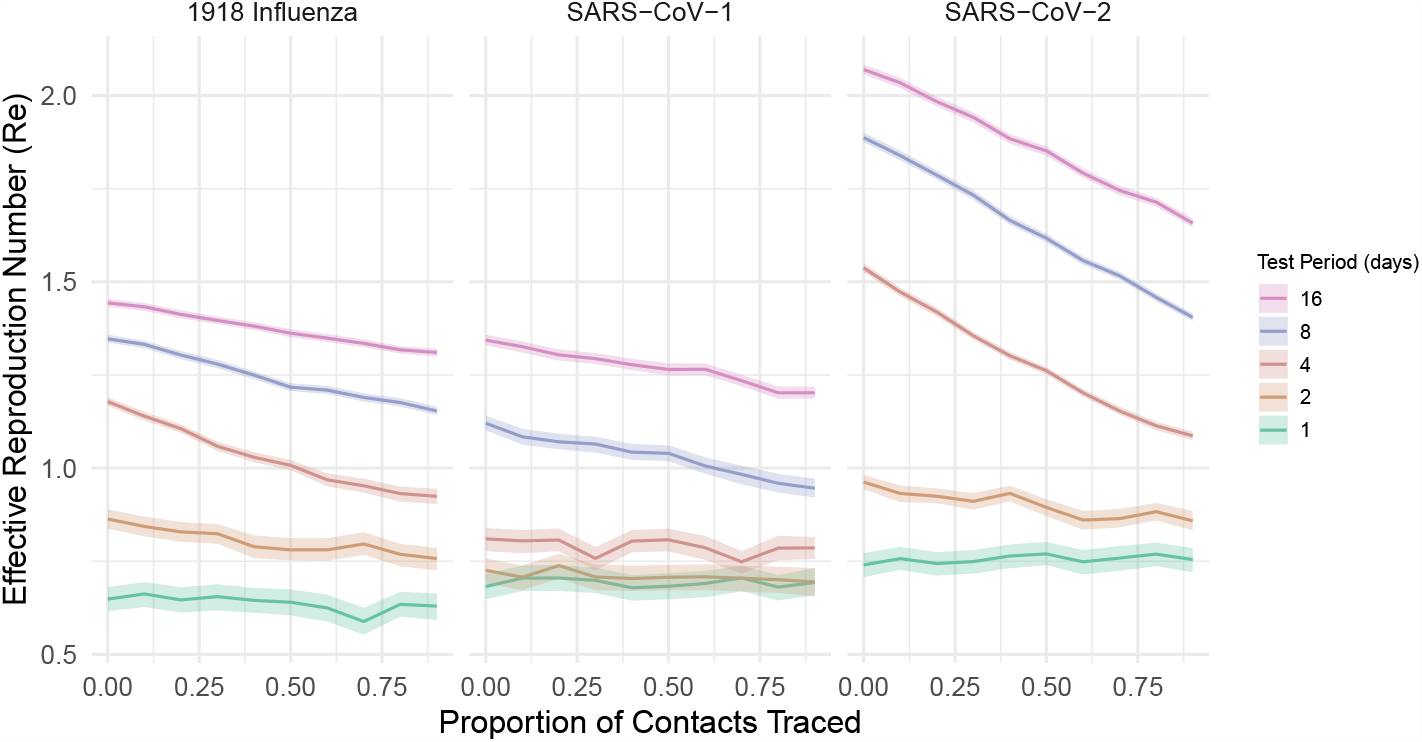
*R*_*e*_ Depending on Testing Period and Proportion of Contacts Traced with Probability a Person Tests After Noticing Symptoms = 0.50. *R*_*e*_ for the 1918 Influenza pandemic (left) is computed using pathogen-specific parameters shown in Table 1 (excluding probability testing once symptomatic), a contact tracing delay of 24 hours, adherence to isolation of 0.90, and the probability a person gets tested after being symptomatic is 0.50 (note that the probability of developing symptoms for Influenza is 0.693, SARS-CoV-1 is 0.925, and SARS-CoV-2 is 0.800). Similarly, the *R*_*e*_ for SARS-CoV-1 is shown (middle) and SARS-CoV-2 (right). Both plots show various trends as a result of variations in testing period.

**Fig. A3:**
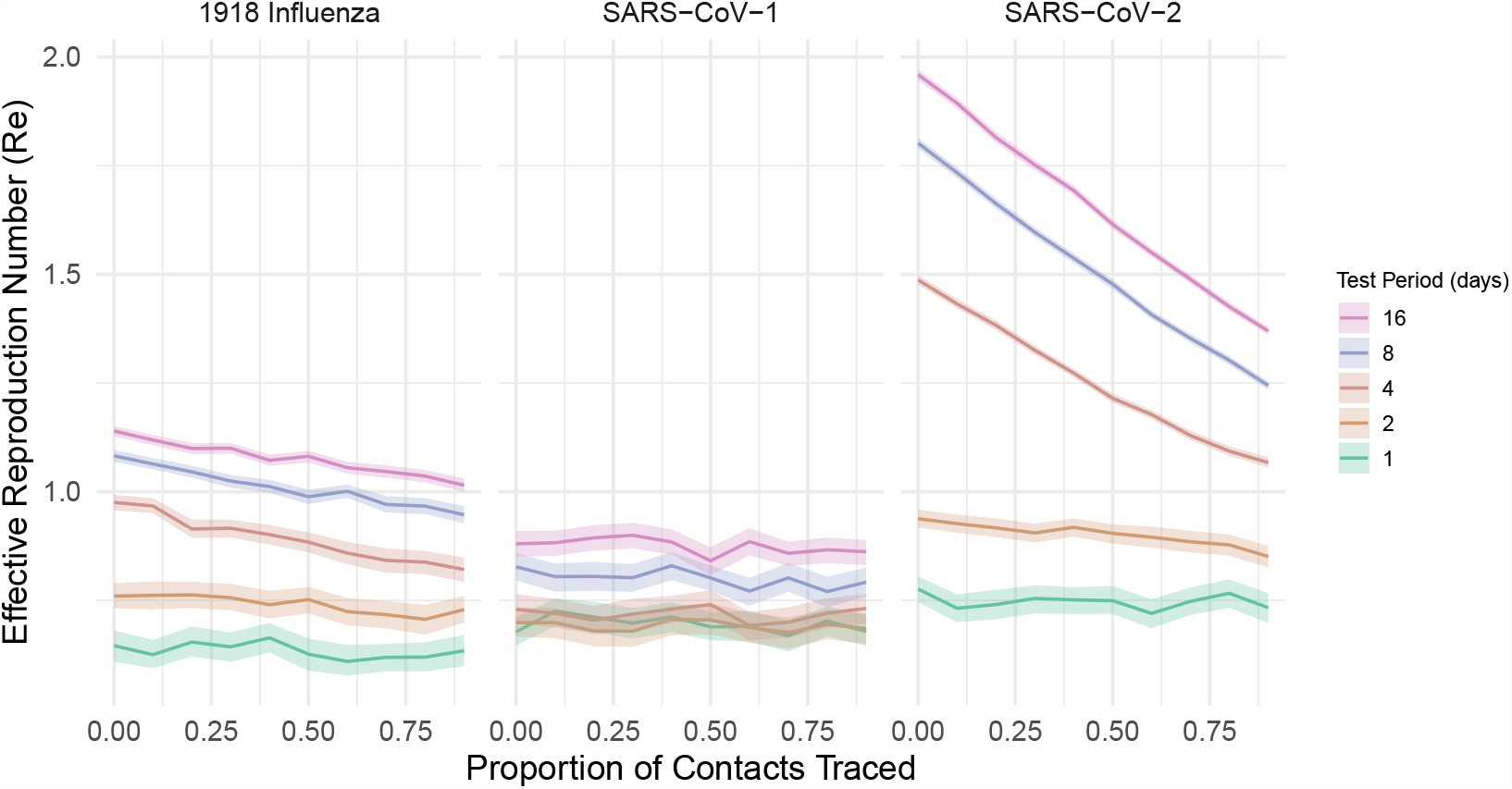
Re Depending on Testing Period and Proportion of Contacts Traced with Probability a Person Tests After Noticing Symptoms = 0.90. The *R*_*e*_ for the 1918 Influenza pandemic (left) is created using pathogen-specific characteristics shown in Table 1 (excluding probability testing once symptomatic), a contact tracing delay of 24 hours, adherence to isolation of 0.90, and the probability a person gets tested after being symptomatic is 0.90 (note that the probability of developing symptoms for Influenza is 0.693, SARS-CoV-1 is 0.925, and SARS-CoV-2 is 0.800). Similarly, the *R*_*e*_ for SARS-CoV-1 is shown (middle) and SARS-CoV-2 (right). Both plots show various trends as a result of variations in testing period.

**Fig. B4:**
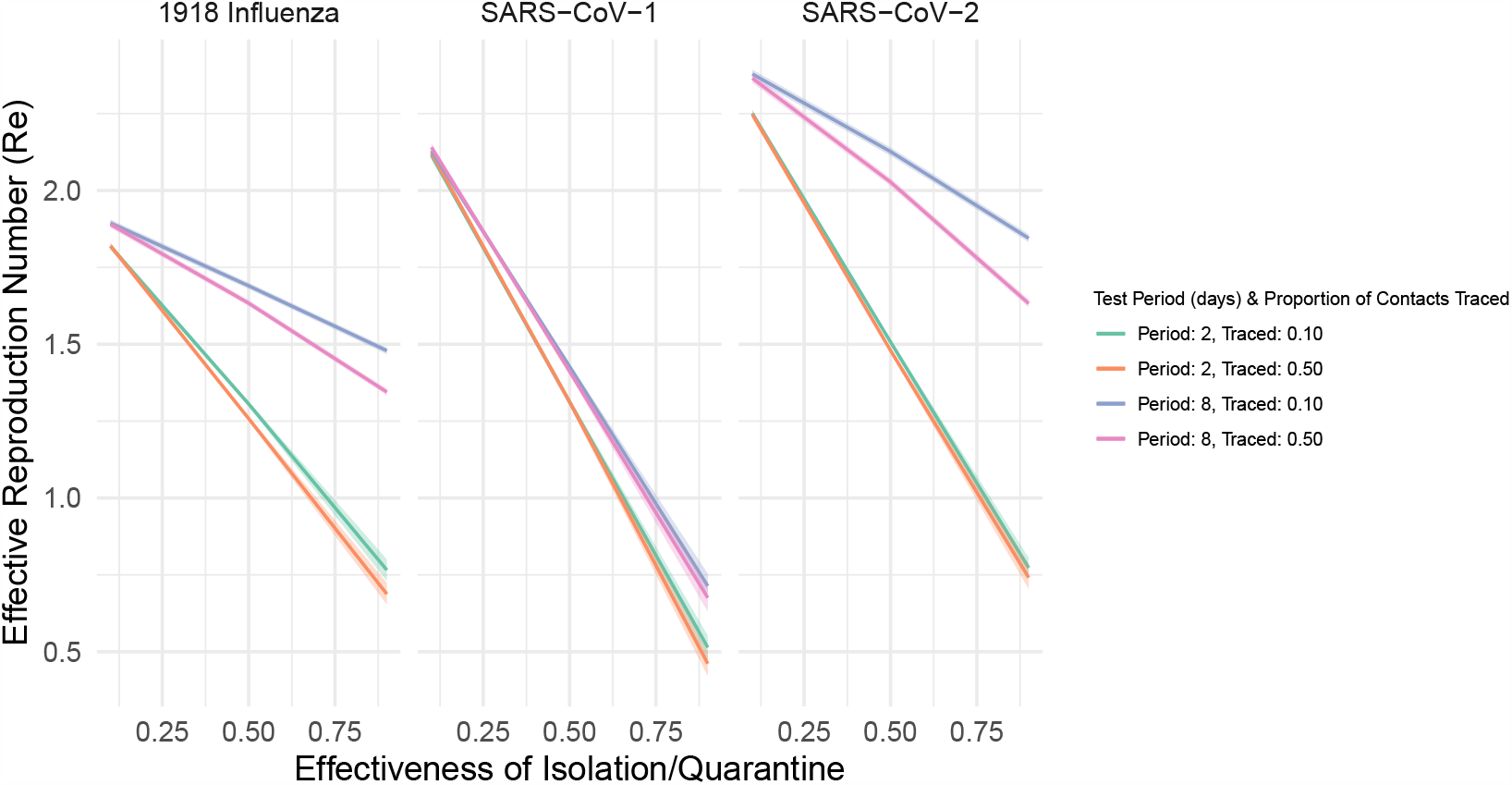
Re Depending on the Effectiveness of Isolation or Quarantine and Testing Period and Proportion of Contacts Traced. The *R*_*e*_ for the 1918 Influenza pandemic (left) is calculated depending on variations in adherence to isolation and using pathogen-specific characteristics shown in Table 1 and a contact tracing delay of 24 hours. Similarly, the *R*_*e*_ for SARS-CoV-1,(middle), and SARS-CoV-2 (right) is shown as a result of variations in the effectiveness of isolation and quarantine. Both plots show various trends as a result of variations in contact tracing and testing period.

## Notes

### Competing Interest Statement

The authors have declared no competing interest.

### Funding Statement

This study did not receive any funding

